# An Ontology-based Approach to Guide and Document Variable and Data Source Selection and Data Integration Process to Support Integrative Data Analysis in Cancer Outcomes Research

**DOI:** 10.1101/2020.05.28.20115907

**Authors:** Hansi Zhang, Yi Guo, Jiang Bian

**Affiliations:** Department of Health Outcomes and Biomedical Informatics, College of Medicine, University of Florida, Gainesville, Florida, USA

**Author notes:** Email: Hansi Zhang –; Yi Guo –; Jiang Bian –. Corresponding author: Jiang Bian,. Department of Health Outcomes and Biomedical Informatics, College of Medicine, University of Florida, 2197 Mowry Road, Suite 122, PO Box 100177, Gainesville, FL 32610-0177.

**Keywords:** Ontology, Integrative data analysis, Cancer outcomes research, Reporting guideline

## Abstract

**Background:** To reduce cancer mortality and improve cancer outcomes, it is critical to understand the various cancer risk factors (RFs) across different domains (e.g., genetic, environmental, and behavioral risk factors) and levels (e.g., individual, interpersonal, and community levels). However, prior research on RFs of cancer outcomes, has primarily focused on individual level RFs due to the lack of integrated datasets that contain multi-level, multi-domain RFs. Further, the lack of a consensus and proper guidance on systematically identify RFs also increase the difficulty of RF selection from heterogenous data sources in a multi-level integrative data analysis (mIDA) study. More importantly, as mIDA studies require integrating heterogenous data sources, the data integration processes in the limited number of existing mIDA studies are inconsistently performed and poorly documented, and thus threatening transparency and reproducibility.

**Methods:** Informed by the National Institute on Minority Health and Health Disparities (NIMHD) research framework, we (1) reviewed existing reporting guidelines from the Enhancing the QUAlity and Transparency Of health Research (EQUATOR) network and (2) developed a theory-driven reporting guideline to guide the RF variable selection, data source selection, and data integration process. Then, we developed an ontology to standardize the documentation of the RF selection and data integration process in mIDA studies.

**Results:** We summarized the review results and created a reporting guideline—ATTEST—for reporting the variable selection and data source selection and integration process. We provided an ATTEST check list to help researchers to annotate and clearly document each step of their mIDA studies to ensure the transparency and reproducibility. We used the ATTEST to report two mIDA case studies and further transformed annotation results into sematic triples, so that the relationships among variables, data sources and integration processes are explicitly standardized and modeled using the classes and properties from OD-ATTEST.

**Conclusion:** Our ontology-based reporting guideline solves some key challenges in current mIDA studies for cancer outcomes research, through providing (1) a theory-driven guidance for multi-level and multi-domain RF variable and data source selection; and (2) a standardized documentation of the data selection and integration processes powered by an ontology, thus a way to enable sharing of mIDA study reports among researchers.

## BACKGROUND

Cancer is a major disease burden worldwide [1]. As the 2nd leading cause of death in the United States (US), about 1 in 4 deaths is due to various types of cancer [2]. In 2019, an estimation of 1,762,450 new cancer cases diagnosed and 606,880 cancer deaths is reported by the American Cancer Society (ACS) in US [2]. The lifetime probabilities of being diagnosed with cancer are 39.3% and 37.4% for male and female, respectively [3]. However, the risk factors for these high cancer incidence and mortality rates are still not fully understood.

Nevertheless, to reduce cancer mortality rates and improve cancer outcomes (e.g., survival and prognosis), it is critical to understand the various risk factors of cancer. So far, evidence suggests that it is the interaction among many risk factors (RFs) together that affect the risk of cancer and cancer outcomes, rather than a single cause [4]. Further, the RFs involved are across different domains (e.g., genetic, environmental, and behavioral risk factors) and levels (e.g., individual level, interpersonal level, and community level). However, there is not yet an agreement among the cancer research community regarding how these multi-level cancer RFs interact with each other. To do so, the first and most crucial step is to gain a comprehensive view of potential multi-level RFs associated with various cancer outcomes such as the stage of diagnosis (the most important prognostic factor) and survival.

We surveyed existing research on RFs for late stage cancer diagnosis and poor survival, we found current studies about RFs for cancer outcomes are mostly from single-level analyses with mostly individual patient-level data. For instance, Andrew *et al*. assessed individual patient characteristics (e.g., age, gender, family history), and lifestyle factors (e.g., education, insurance and socioeconomic status) to study their risks associated with colorectal cancer at late stage [5]. These individual-level RFs have also been reported for other major types of cancers such as breast and cervical cancers [6–9]. Further, prior studies studying cancer RFs often only analyzed data from a single source, such as SEER [10], SEER-Medicare[11], or a state or hospital cancer registry [12]. Among these cancer risk factor studies, the complex interplay between difference levels RFs are often ignored (e.g., county-level smoking rate vs. individual smoking behavior). These single-level RF analyses (1) lead to biased effect estimates of RFs due to potential confounding from omitted factors, (2) omit critical cross-level RF interactions, such as race by residence, that could inform multi-level intervention design.

Nowadays, advances in technology created new ways for us to determine and measure disease risk factors across different levels (e.g., from advancements in genome sequencing for genetic markers to better sensors for producing more accurate estimates of environmental pollutants). The availability of such abundant data online in electronic formats enables researchers to pool data on an unprecedented scale and offers a great opportunity to do a thorough examination of multi-level RFs in a multi-level integrative data analysis (mIDA) so that confounding effects and across-level interactions can be studied. However, researchers face significant barriers to do so, especially because there is a lack of consensus and proper guidance to help researchers systematically think and discovery these variables from heterogenous sources. In 2017, National Institute on Minority Health and Health Disparities (NIMHD) of the National Institute of Health (NIH) proposed a Research Framework [13], an extension to the well-known social ecological model [14], to help investigators systematically study health disparities. Recognized by the NIMHD Framework, individuals are embedded within the larger social system and constrained by the physical environment they live in. Within this framework, cancer outcomes are influenced by RFs from different levels (i.e., individual, interpersonal, community, and societal) and multiple domains (i.e., biological, behavioral, physical/built environment, sociocultural environment, and healthcare system). In this work, we adopted the NIMHD framework as the guiding theory for risk factor discovery and data source selection.

Further, mIDA for cancer outcomes research requires the integration of data from multiple sources. However, data integration processes in the very limited number of existing mIDA studies [15, 16] are inconsistently performed and poorly documented, and thus threatening transparency and reproducibility [17, 18]. The data integration processes are often time summarized in one or two sentences without explicitly documentation of the steps. For example, Guo *et al*. explored the impact of the relationships among socioeconomic status, individual smoking status, and community-level smoking rate on pharyngeal cancer survival [16]. The multi-level risk factors above were obtained and integrated from three different data sources (i.e., Florida Cancer Data System [FCDS], U.S. Census, and Behavioral Risk Factor Surveillance System [BRFSS]) as mentioned in the abstract. However, for the rest of the paper, there is no description of how the individual-level records from FCDS are linked with county-level smoking rate from BRFSS and census tract-level poverty rate from U.S. Census. Even though the integration process might be as simple as integrating these multi-level variables through the geographic code (e.g., county code), it still needs to be standardized and explicitly documented to avoid ambiguity. For example, the paper discussed that “*regional smoking was measured as the average percentage of adult current smokers at the county level between 1996 and 2010*” and the readers might be able to make an educated guess that the regional smoking rates were more likely to be generated using the BRFSS data rather than from the FCDS data; however, explicit documentation is needed as both BRFSS and FCDS data have individual smoking status. Keegan *et al*. explored and whether breast cancer survival patterns are influenced by factors such as nativity (individual level) and neighborhood socioeconomic status (community level). Similarly, they summarized integration process in one sentence by stating each patient was assigned a neighborhood socioeconomic status variable based the census block groups. However, the details such as variable names in each data sources, or whether the original geographic variables require pre-processing (e.g., derive census tract from zip codes) are not clearly documented [15]. The explicit documentation of these variable selection and data integration processes will help readers to better understand the study results, benefit other researchers who want to replicate the studies, but also more importantly, make it possible for machines to understand and replicate the steps (when these explicit documentations are encoded in a computable format such as with an ontology).

Further, even though these mIDA studies above did not emphasize the need for data integration or integrated datasets, the fact that they can only investigated a handful of variables at a time indicated the lack of but needed support on data integration. Even in studies on building frameworks or platforms to support or automate the data integration process (especially those related to creating integrated dataset to support cancer research), they often ignored the need for documenting the integration steps to guarantee the transparency and reproducibility of their approaches. For example, semantic data integration approach —connecting variables across different databases at the semantic level through mapping them to standardized concepts in a global schema (e.g., often time a global ontology) — has been proposed in data integration studies in recent years to support generating integrated datasets for cancer research [19–21]. However, none of these studies mentioned the need for standardizing and documenting their integration steps, for example, most of them did not even discuss the rationale for selecting the specific data sources to integrate. Nevertheless, when reporting mIDA studies, it is critical to document the steps that were followed to select, integrate, and process the data so that others can repeat the same steps and reproduce the findings.

To address challenges above, in this paper, we first developed a reporting guideline to guide and document the RF variable selection, data source selection, and data integration process. The guideline is informed by (1) the NIMHD research framework that provides guidance and promotes structural thinking on identifying multi-level cancer RFs; and (2) reviewing existing reporting guidelines from the Enhancing the QUAlity and Transparency Of health Research (EQUATOR) network [22]. Then, we proposed an ontology-based approach to annotate and document the RF selection and data integration process in mIDA studies based on the reporting guideline we developed. To do so, we developed the **<u>O</u>**ntology for the **<u>D</u>**ocumentation of v**<u>A</u>**riable selec**<u>T</u>**ion and da**<u>T</u>**a sourc**<u>E S</u>**election and in**<u>T</u>**egration proess (OD-ATTEST) so that the RF selection and data integration report can be (1) explicitly modeled with a shared, controlled vocabulary, (2) understandable to humans and computable to computers, and (3) adaptive to changes when the reporting process is refined.

In our prior work [23], we proposed a preliminary reporting guideline for RF variable and data source selection based on our own experience of pooling multi-level RFs from different data sources to support mIDAs of cancer survival [24, 25]. In this extended journal paper, we significantly expanded our ontology-based reporting guideline—ATTEST (v**<u>A</u>**riable selec**<u>T</u>**ion and da**<u>T</u>**a sourc**<u>E S</u>**election and in**<u>T</u>**egration):

- We conducted a systematic search of existing reporting guidelines from the EQUATOR network to extract reporting elements relevant to variable selection and data integration.
- We updated our reporting guideline based on the result of the systematic review to include new items regarding data integration (e.g., data processing, data integration strategy, data validation, etc.) as well as variable and data source selection.
- We completed building the OD-ATTEST following the best practice in ontology development to provide a formal presentation for the reporting guideline with standardized and controlled vocabularies.
- We provided an ontology (OD-ATTEST) annotated report generated based on a prior mIDA study to represent the annotated items and their relationships in reporting guideline.

## METHODS

### Development of a reporting guideline for risk factor selection, data source selection, and data integration

To develop the reporting guideline, we started with summarizing our previous studies where we assessed the effect of data integration on predictive ability of cancer survival models [24] and created a semantic data integration framework to pool multi-level RFs from heterogenous data sources to support mIDA [25]. In the above studies, we went through the process of RF selection, data source selection, and data integration. To be able to ensure the reproducibility of these studies, a number of middle steps need to be documented as detailed in our previous paper [23]. For example, both rural-urban commuting area (RUCA) codes [26] and the National Center for Health Statistics (NCHS) urban-rural classification scheme [27] are often used to represent an geographic area’s rurality status. The difference between the two resides in the classification granularity, where RUCA focuses on classifying U.S. census tracts (i.e., tens levels from rural to metropolitan) while the NCHS urban-rural classification scheme focuses on classifying U.S. counties (i.e., a hierarchal definition with six levels). Thus, we need to clearly document which rural definition we used in the data analysis since different representations of the same variable (i.e., rurality in this case) have different impacts on model results, as shown in our prior work [24]. Further, before integration RFs from various data sources at different levels (e.g., census tract level vs. county level) and covered different time periods, we made an assumption assumed that area-level characteristics (e.g., social vulnerability index) derived from 2000 U.S. Census data were applicable across different time periods (as our individual level data from FCDS covered 1996 and 2010). Above experiences suggest that we must document these data integration nuances so that other researchers can repeat our data integration and data processing pipeline and reproduce the same results (e.g., integrated dataset). In sum, we summarized 3 key items that need to be documented: (1) RF selection (e.g., individual vs county-level variables), (2) data source selection (e.g., individual-level data from FCDS and contextual-level data from US Census), and (3) data integration and data preprocessing strategies.

Through discussions with expert biostatisticians, data analysts, and cancer outcomes researchers, we summarized the typical mIDA process and found there is little structured thinking when investigators selecting and identifying risk factors and their data sources. We thus propose to use the NIMHD research framework to provide a theory-driven guidance for multi-level and multi-domain RF and data source selections. The NIMHD framework is originally designed to depicts a wide range of health determinants (i.e., RFs from different levels and domains) relevant to understanding and addressing minority health and health disparities. The goal of using the NIMHD framework is to help investigators to structurally and comprehensively think and identify relevant RFs and corresponding data sources in their IDA studies.

To build upon existing established reporting guidelines, we searched and identified relevant reporting guidelines from the Enhancing the QUAlity and Transparency Of health Research (EQUATOR) network—a comprehensive searchable database of guidelines for health research reporting. The EQUATOR network categorizes health researches into 13 study types (e.g., quantitative studies, experimental studies, and observational studies), where reporting guidelines for observational studies are most relevant to our mIDA use case. To further identify relevant reporting guidelines in EQUATOR, we developed a set of screening criteria to determine whether a reporting guideline in EQUATOR contains the information that can be used to improve our ATTEST reporting guideline as shown below:

- The reporting guideline is designed for secondary data analysis studies.
- The reporting guideline contains at least one of the following sections: data, outcomes (variables), and methods, as these sections will contain information related to variable selection, data source selection, and data integration methods.
- The reported data within the guideline must be health related.
- The use of the guideline (at least part of the guideline) can be extended to the cancer outcomes research, especially those related to variable selection, data source selection, and data integration.

We reviewed all reporting guidelines designed for observational studies and eliminated guidelines that do not involve the tasks of RF and data source selection and integration. We then identified all reporting guidelines that contain the following sections: data, outcomes (variables), and methods. For those that do not have sections clearly marked, we manually reviewed the entire reporting guideline to identify whether they discussed one of the three aspects. We then extracted reporting items in the selected reporting guidelines that are relevant to RF selection, data source selection, and data integration. Two reviewers (HZ and JB) independently extracted these reporting items of interest and resolved conflicts with a third reviewer (YG). We further analyzed these extracted reporting items and discussed with experts (i.e., biostatisticians, data analysts and cancer outcomes researchers) to summarize items needed in our reporting guideline, especially those related to the data integration process.

### Construction of an ontology for the documentation of variable and data source selection and integration process (OD-ATTEST)

The ATTEST reporting guideline we developed is used to guide the variable and data source selection and integration process in cancer outcomes research. We propose to use an ontology-based approach to annotate and document the items in the reporting guideline. The goal of the OD-ATTEST ontology is to standardize the terminology used in documenting the selection and integration steps of RF variables and data sources to support mIDA.

The OD- ATTEST is developed using Protégé 5. We used Basic Formal Ontology (BFO) [28] as the upper-level ontology. We first adopted a top down approach to enumerate important entities (classes and relations) based on the reporting guideline we developed. Following the best practice, we reviewed existing widely accepted ontologies using the National Center for Biomedical Ontology (NCBO) BioPortal [29] to find the entities can be reused in OD-ATTEST. Then, we started with the definitions of the most general concepts in the domain and subsequent specialization of the concepts to develop the class hierarchy. We also took a bottom-up process, where we started with the definitions of the most specific classes, and then subsequent grouped similar classes into more general concepts. We also examined how these reporting items are associated with each other (e.g., “*sample size*” is determined by “*primary outcome*”) and determined what additional classes and relations were needed to fully represent these entities in OD-ATTEST.

### An OD-ATTEST-annotated report generated based on a mIDA case study following the reporting guideline

To test the developed ATTEST reporting guideline and the OD-ATTEST ontology, we first created a ATTEST report based on our previous mIDA case study, where we explored the impact of the relationships among socioeconomic status, individual smoking status, and community-level smoking rate on pharyngeal cancer survival [16]. To annotate the ATTEST report using OD-ATTEST, we used the following annotation process: 1) identify information related to the reporting items in ATTEST through reviewing the original publication and supplementary materials; 2) annotate the information using the entities in OD-ATTEST; and 3) transform annotation results into semantic triples in Resource Description Framework (RDF) format using Turtle syntax [30].

## RESULTS

### The ATTEST reporting guideline for RF variable and data source selection and data integration

We extended our preliminary reporting guideline [23] through a review of existing relevant reporting guidelines published in the EQUATOR network. ***Figure 1*** shows our review process. We reviewed 94 reporting guidelines designed for from observational studies in the EQUATOR network. Out of the 94 reporting guidelines, 30 contain the required data, outcomes (variables), and method sections, which we retained for data extraction. In the data extraction step, for each reporting guideline, we extracted items relevant to RF and data source selection and integration, where the data and outcomes (variables) sections often contain information regarding how RF variables and data sources are selected, while the method section contains information about how data are processed and integrated.

**Figure 1.**
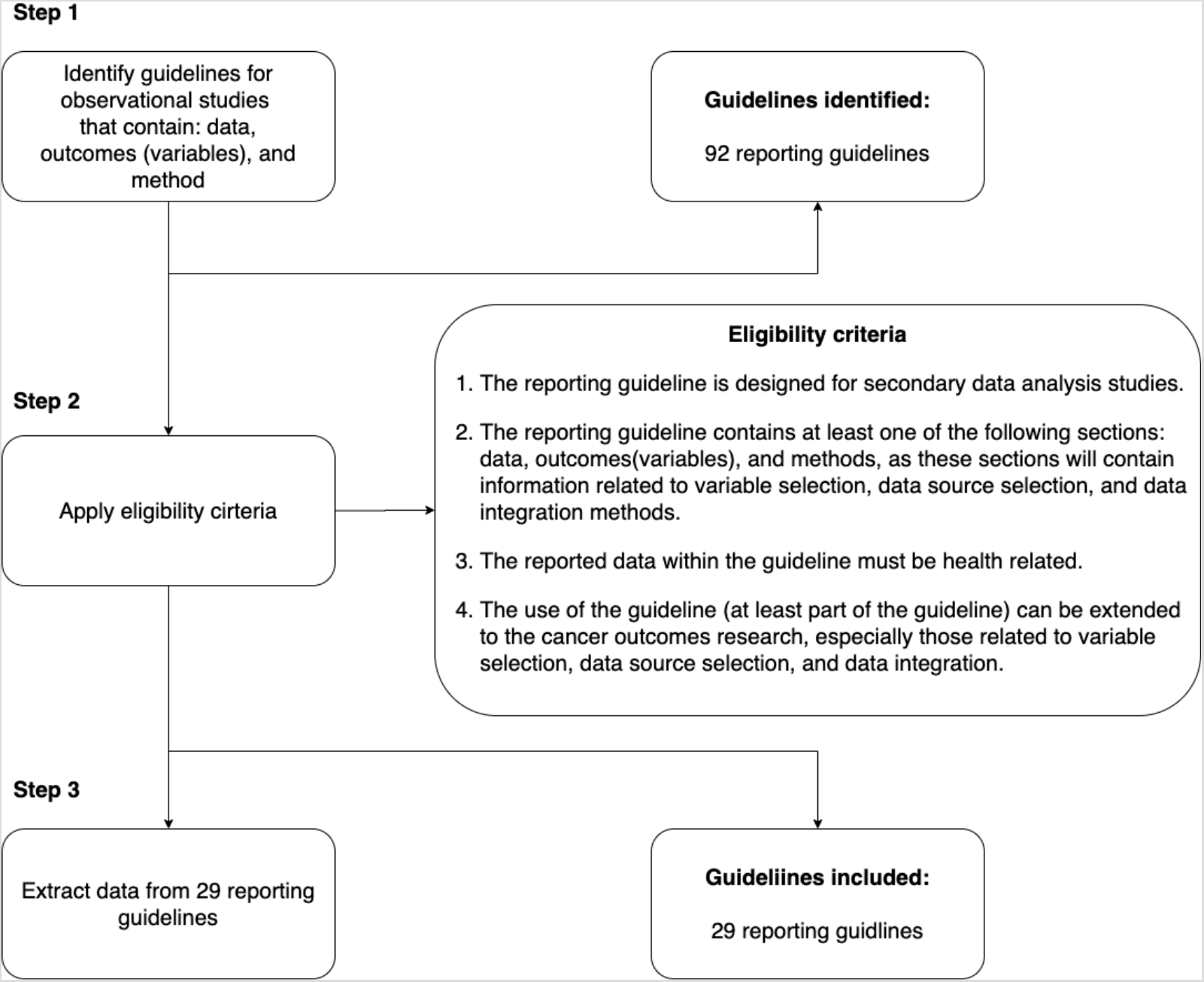
Review of relevant reporting guidelines in the EQUATOR network.

We categorized these reporting guidelines (***Table 1***) based on the domains and levels of the data sources reported in the guidelines and mapped them to the NIMHD framework. As shown in ***Table 1***, these 29 reporting guidelines cover data sources from all domains and levels of influences. Among them, 9 guidelines focused on providing a general reporting guideline for observational studies without specifying a specific domain of influence; while the rest of the guidelines are designed for different domains. For example, the Genetic RIsk Prediction Studies (GRIPS) statement [31] is designed for risk prediction studies using genetic data. Furthermore, most guidelines only considered the data sources from individual level, while 2 of them considered data sources from multi-levels. For example, the Checklist for One Health Epidemiological Reporting of Evidence (COHERE) [32] considered both individual and environmental risk factors when studying a disease.

**Table 1.**
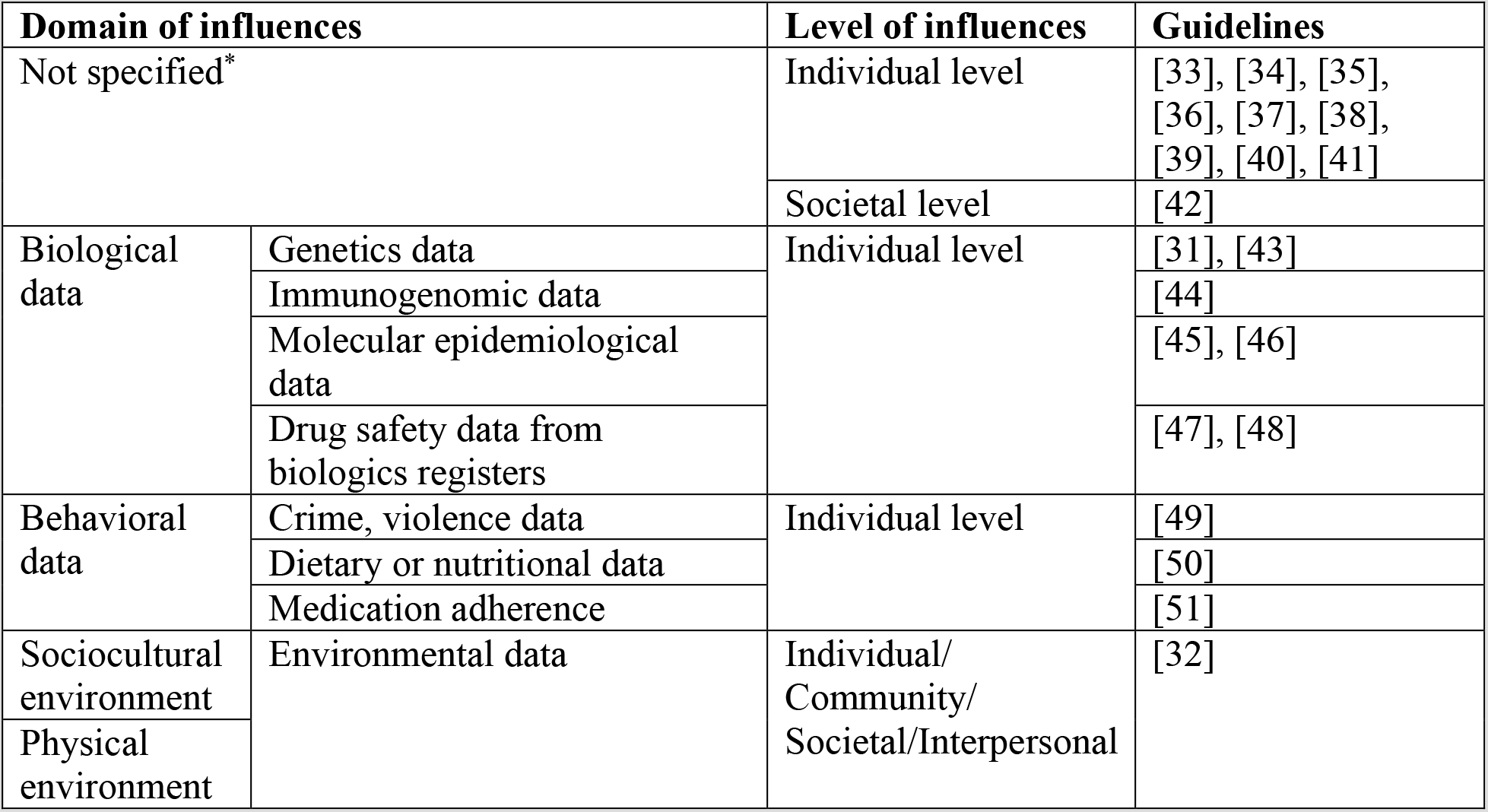
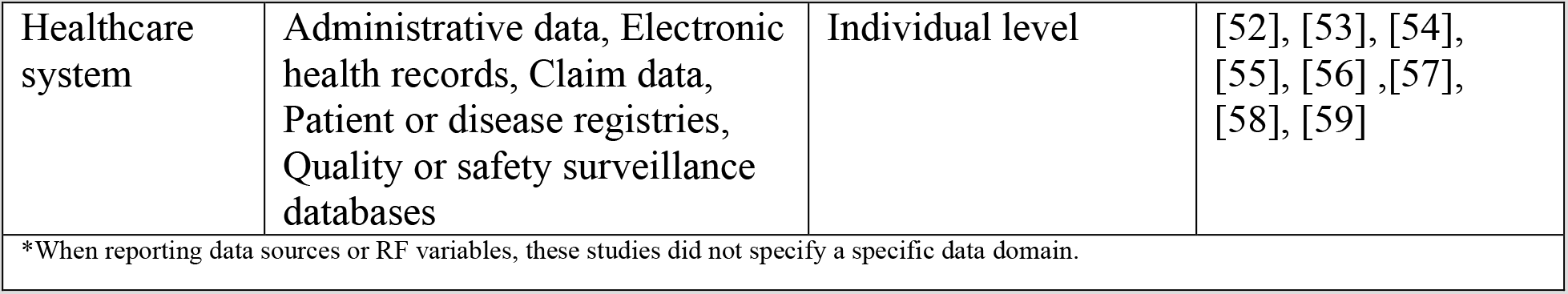
Summary of reporting guidelines based on the data source domains and levels guided by the NIMHD framework.

In our preliminary reporting guideline [23], we focused only on reporting items relevant to RF variables and data sources selection. In this review, we extracted items that can be used to improve our initial reporting guideline but with a focus on documenting the data integration process. In total, we found 3 reporting guidelines [53–55] contain information about data integration processes. However, items included in these 3 guidelines focus on data linkage and do not contain enough details about how to solve the heterogeneities of data from different sources. For example, when integrating variables across different levels (e.g., combine individual-level patient data and county-level smoking rate), none of the 3 guidelines have items on documenting the cross-level integration choices (e.g., layering the county-level smoking rate to individual based on residence of the individuals and county code), while this type of choices is frequently encountered in mIDA studies. Further, data processing steps such as the choices and algorithms used for creating new data elements (e.g., compute a body mass index variable from two separate variables, weight and height) are not documented in existing reporting guidelines. Therefore, we further extended the ATTEST to include these important data integration and data processing procedures based on our previous research experience on building data integration framework [25].

Informed by the NIMHD research framework and consistent with our prior work, the ATTEST reporting guideline consists of two main parts as shown in ***Figure 2***, reporting (1) the objective of the study including explaining the background and rationale for designing the study in one or two sentences and describing the hypothesis of the study; and (2) the study design for variable and data source selection processes and describing the data along with the data integration and processing strategies. The variable and data source selection process consist of five key steps: (1) define the outcome variables for primary and (if necessary) secondary outcomes; (2) for each outcome variable, follow an iterative process (see ***Figure 2.A***) to determine the data sources according to NIMHD framework. After selecting each outcome variable and data sources, investigators need to think about how to select or consolidate similar outcome variables from the different selected data sources. For example, if the outcome of interest is an individual’s lung cancer risk, we shall first identify potential data sources (e.g., cancer registries or electronic health records [EHRs]) that contain individual-level patient data where lung cancer incidence data are available. Then, based on the cohort criteria and other information such as required sample size and data range (e.g., time coverage and geographic information) of the potential data sources, the investigator could determine the qualified data sources and choose an adequate one based on the objective and design of the study. For example, if 2 data sources, cancer registry and EHRs, are both available and contain individual-level lung cancer incidence data, the investigator has the choices to (1) choose one data source over the other, or (2) link the two data sources and integrate variables from the two data sources. If the investigator chooses to link and integrate the two data sources, she needs to explicitly document the linkage and integration processes for each of variables as shown in ***Figure 2*** (Report – Variables – E, F, G, H) so that others can repeat the processes to generate the same analytical dataset; (3) determine the individual-level predictors and covariates of the study; (4) for each individual-level predictor or covariate, follow loop B in ***Figure 2*** to identify the different levels/domains of predictors or covariates according to NIMHD framework. Similar to the outcome variables, different data sources could potentially contain the same predictor or covariate variable, thus, it is important to contrast and consolidate a new predictor or covariate with the existing selected predictors and covariates to resolve duplicates. If an investigator chooses to integrate the “duplicate” variables (e.g., choosing smoking status from cancer registry data over EHRs because cancer registries data are manually abstracted and typically have better data quality than raw EHRs), these data integration choices also need to be explicitly documented. Nevertheless, it is often a difficult choice and these “duplicate” variables might all need to be tested in models before a selection can be made. Regardless, these decisions and data processing steps need to be clearly documented; and (5) after selecting individual-level predictors and covariates, one can use a similar process, following loop C in ***Figure 2*** to identify additional contextual-level predictors and covariates and data sources of interest. In the end, a report of the selected data and data sources as well as the data integration processes shall be generated as shown in ***Figure 2***. The corresponding ATTEST reporting guideline checklist is shown in ***Table 2***.

**Figure 2.**
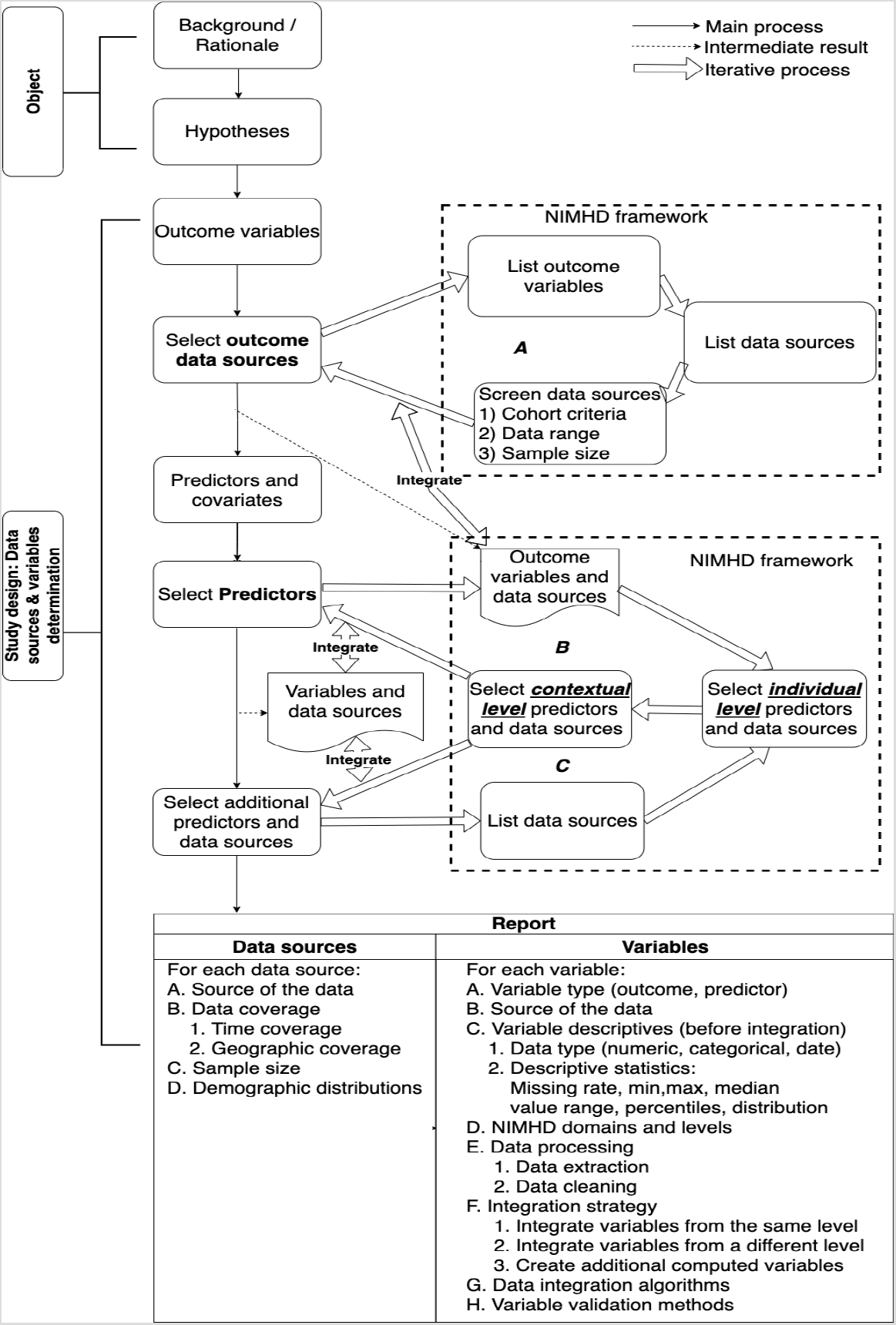
An overview of the reporting guideline for RF variable and data source selection and data integration.

**Table 2.** ATTEST reporting guideline checklist

|  | **Item No** | **Recommendation** | **Page No** |
| --- | --- | --- | --- |
| **Objectives** |  |  |  |
| Background/rationale | 1 | Explain the scientific background and rationale for the study being reported in one or two sentences |  |
| Prespecified hypotheses: | 2 | State prespecified hypotheses in one or two sentences |  |
| **Study design: data sources selection & variables selection & data integration** |  |  |  |
| Data sources | 3a | Describe the time coverage |  |
|  | 3b | Describe the geographic coverage |  |
|  | 3c | Describe the sample size |  |
|  | 3d | Describe the demographic distribution |  |
|  | 3e | Describe the cohort criteria |  |
|  | 3f | Describe the sources of biases (e.g., sample bias) |  |
|  | 3g | Describe the data collection approach |  |
| Dependent variables | 4a | State the variable definition and variable type (e.g., primary outcome variable, secondary outcome variable) |  |
|  | 4b | State the data source of dependent variable |  |
|  | 4c | State the data type (e.g., numerical, categorical, date-time) of dependent variable |  |
|  | 4d | State descriptive statistics (e.g., min, max, median, value range, percentile) of dependent variable |  |
|  | 4e | State the NIMHD domains and levels of dependent variable |  |
| Independent variables | 5a | State the variable definition and variable type (e.g., primary predictor, secondary predictor) |  |
|  | 5b | State the data source of dependent variable |  |
|  | 5c | State the data type (e.g., numerical, categorical, date-time) of dependent variable |  |
|  | 5b | State descriptive statistics (e.g., min, max, median, value range, percentile) of independent variable |  |
|  | 5e | State the NIMHD domains and levels of independent variable |  |
| Controlled variables | 6a | State the variables type (e.g., numerical, categorical) of controlled variable |  |
|  | 6b | State the data source of controlled variable |  |
|  | 6c | State descriptive statistics (e.g., min, max, median, value range, percentile) of controlled variable |  |
|  | 6d | State the NIMHD domains and levels of controlled variable |  |
| Missing data | 7a | For each data source, describe whether required or expected variable that is not present |  |
|  | 7b | For each variable, describe method of how to handle missing data |  |
|  | 7c | For each variable, describe the missing rate |  |
| **Data integration** |  |  |  |
| Data processing | 8a | Data extraction: for each variable, describe how to process the raw data source to extract the variable |  |
|  | 8b | Data cleaning: for each variable, describe the method used to detect and correct (or remove) the incorrect records, missing values or outliers |  |
| Integration strategy | 9 | Describe the integration strategy for each variable: 1) Integrate with variables from same level, 2) Integrate with variables from different levels, and 3) Creation of additional computed elements |  |
| Integration algorithm | 10 | For each variable, describe the algorithm used to integrate it with variables from other data sources |  |
| Variable validation | 11 | For each variable, describe data validation rule for the selected variable. Rule should identify both the variable and the validation algorithms |  |
| Integrated variable | 12 | Describe the variable after integration and basic descriptive statistics (e.g., min, max, median, value range, percentile) |  |

\*National Institute on Minority Health and Health Disparities (NIMHD)
Note: Please document the items for each data source and variable separately.

### Development of the OD-ATTEST ontology

Based on the ATTEST reporting protocol above, we identified that 48 classes and 25 properties are needed in OD-ATTEST to represent the ATTEST reporting guideline. ***Figure 3*** shows the class hierarchy of OD-ATTEST. We reused classes from the following existing well-known ontologies: Ontology for Biomedical Investigations (OBI), Information Artifact Ontology (IAO), National Cancer Institute Thesaurus (NCIt), Statistics Ontology (STATO) and Semanticscience Integrated Ontology (SIO) as shown in ***Table 3***. Note that there are very few existing ontologies designed for the purpose of documenting the variable and data source selection and data integration process. The limited number of properties in these existing ontologies are not informative to represent the elements in the reporting guideline and their relationships, requiring us to create a large number of new properties in **OD-ATTEST**.

**Figure 3.**
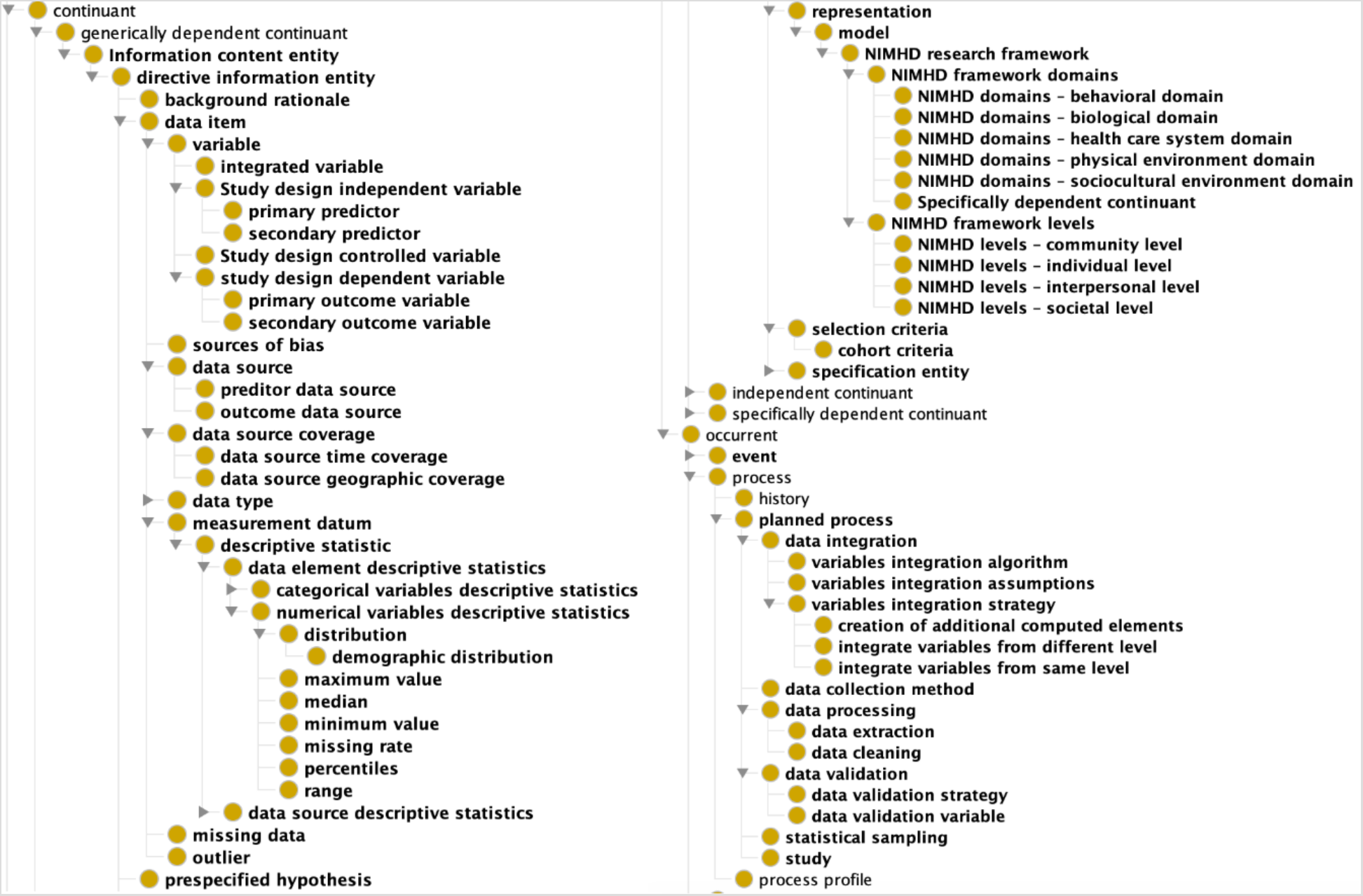
The class hierarchy of OD-ATTEST.

**Table 3.**
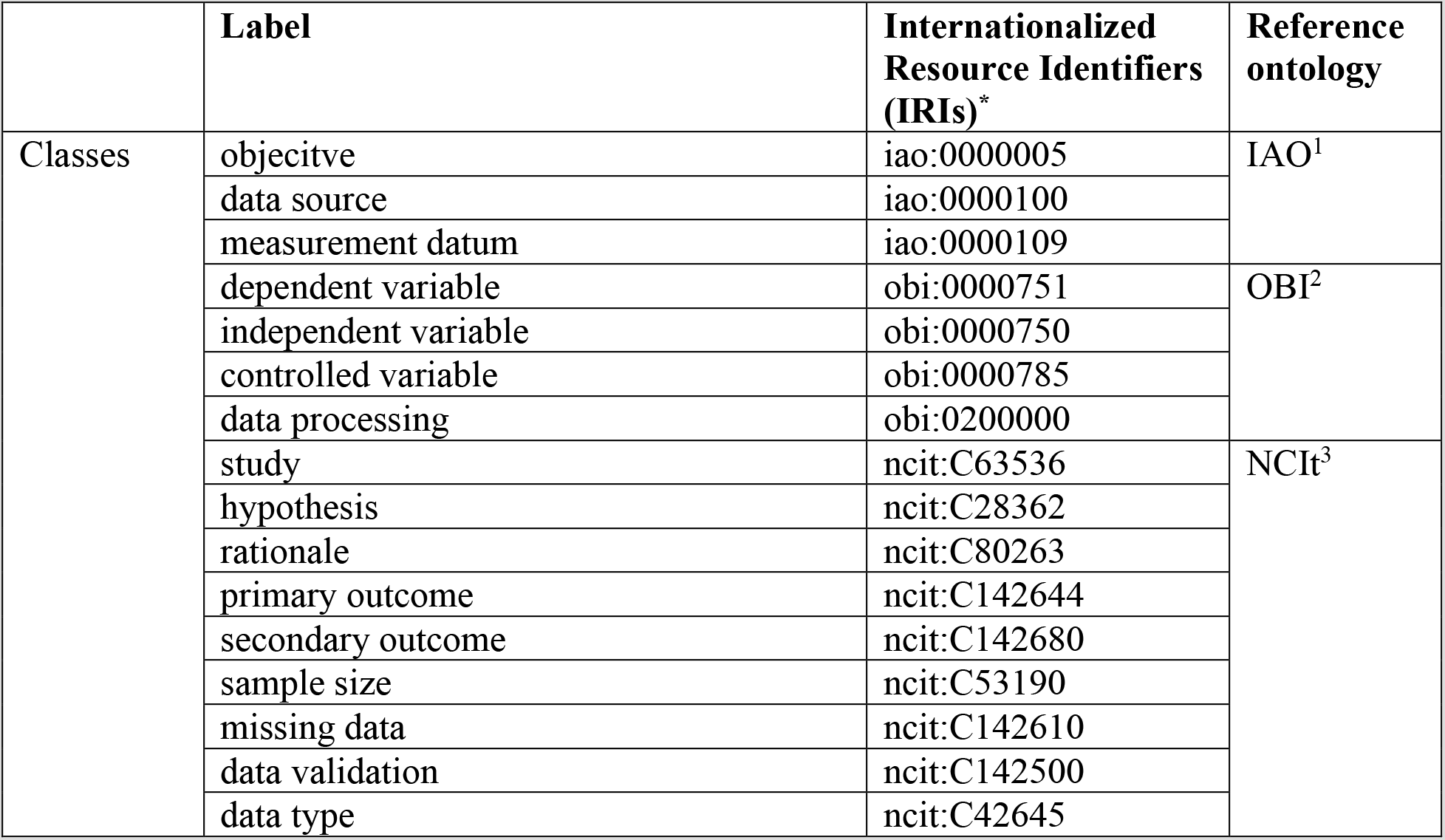
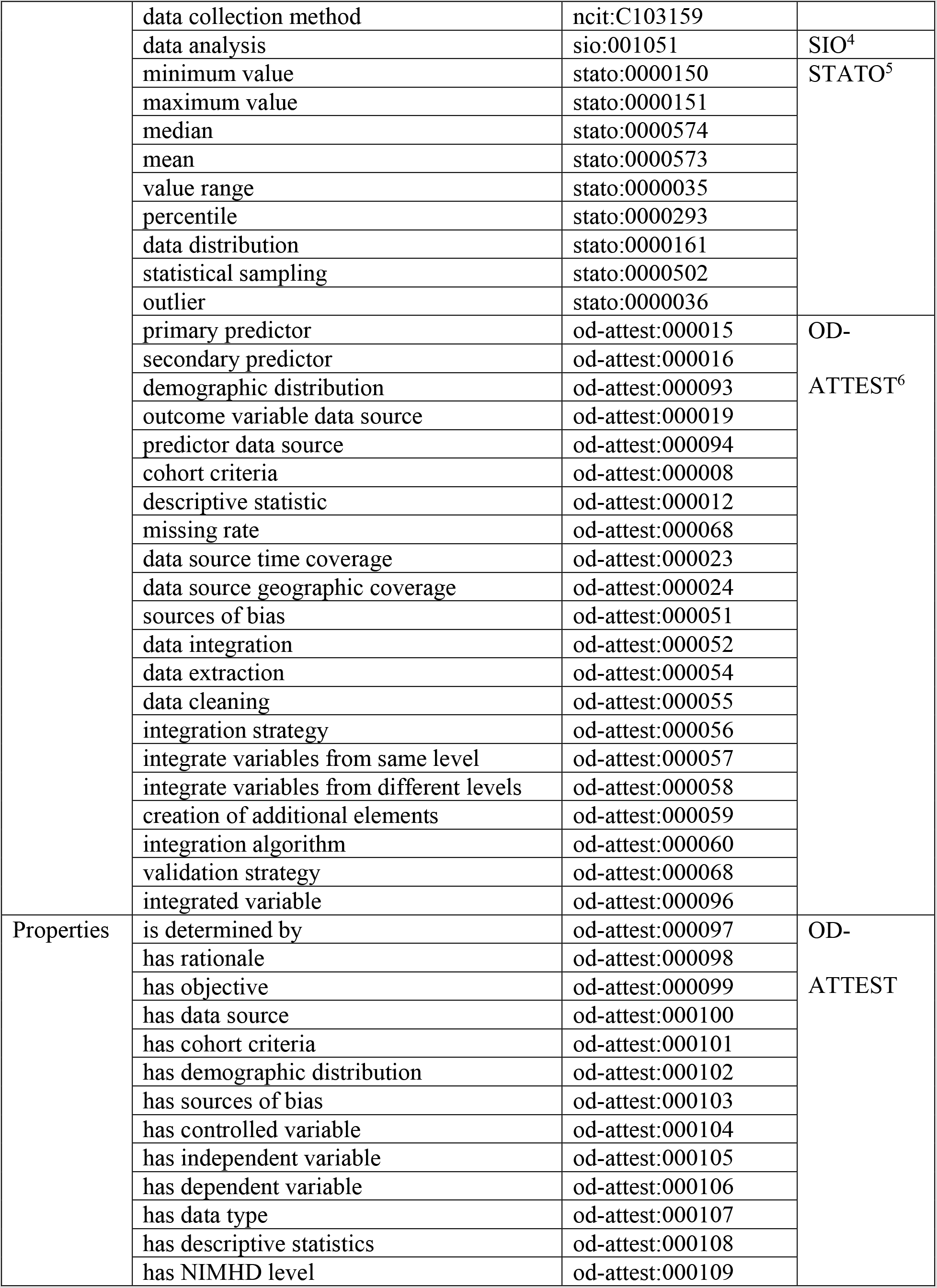
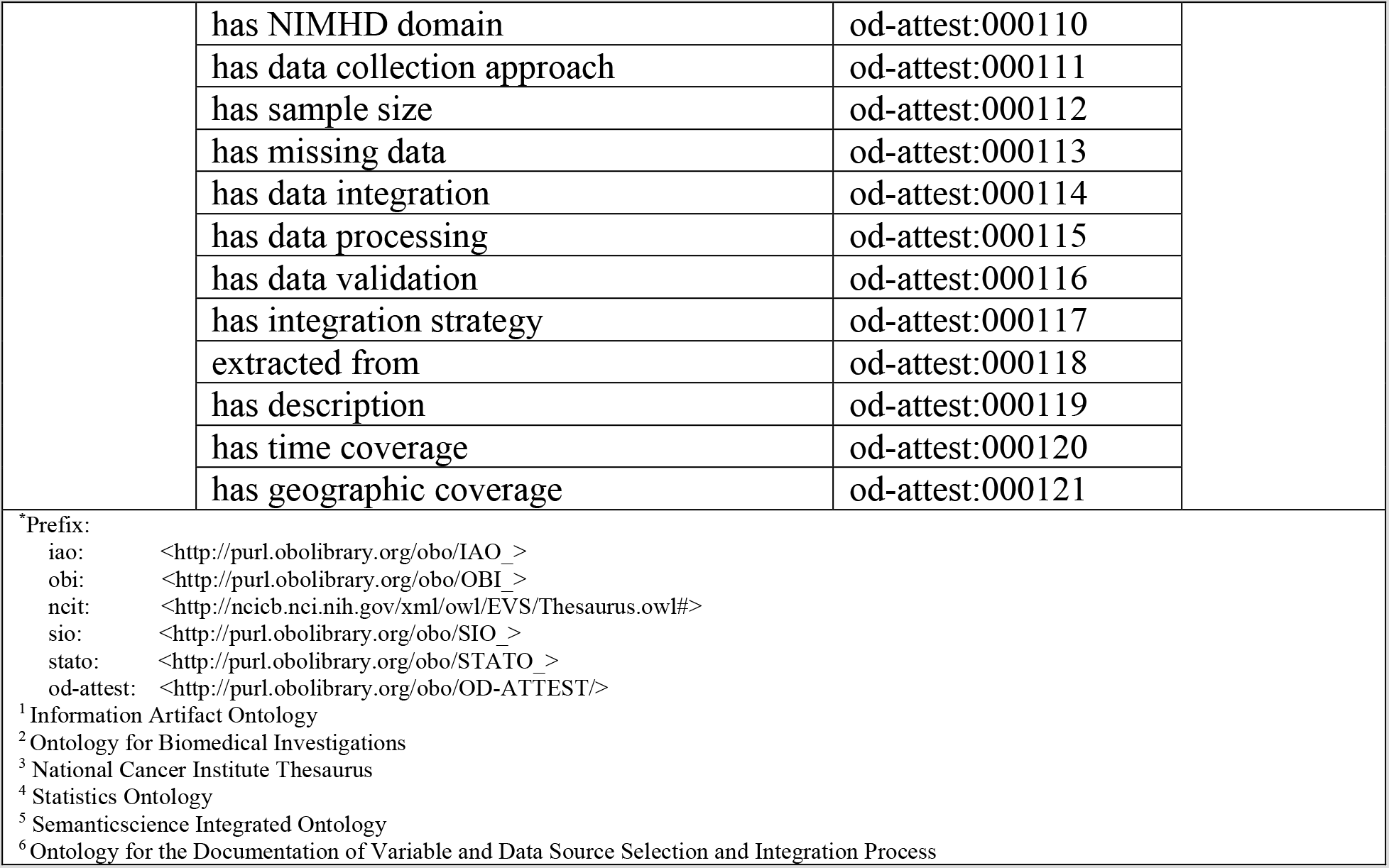
The classes and properties reused or created for OD-ATTEST.

|  | Label | Internationalized Resource Identifiers (IRIs)* | Reference ontology |
| --- | --- | --- | --- |
| Classes | objective | iao:0000005 | IAO<sup>1</sup> |
|  | data source | iao:0000100 |  |
|  | measurement datum | iao:0000109 |  |
|  | dependent variable | obi:0000751 | OBI<sup>2</sup> |
|  | independent variable | obi:0000750 |  |
|  | controlled variable | obi:0000785 |  |
|  | data processing | obi:0200000 |  |
|  | study | ncit:C63536 | NCIt<sup>3</sup> |
|  | hypothesis | ncit:C28362 |  |
|  | rationale | ncit:C80263 |  |
|  | primary outcome | ncit:C142644 |  |
|  | secondary outcome | ncit:C142680 |  |
|  | sample size | ncit:C53190 |  |
|  | missing data | ncit:C142610 |  |
|  | data validation | ncit:C142500 |  |
|  | data type | ncit:C42645 |  |
|  | data collection method | ncit:C103159 |  |
|  | data analysis | sio:001051 | SIO<sup>4</sup> |
|  | minimum value | stato:0000150 | STATO<sup>5</sup> |
|  | maximum value | stato:0000151 |  |
|  | median | stato:0000574 |  |
|  | mean | stato:0000573 |  |
|  | value range | stato:0000035 |  |
|  | percentile | stato:0000293 |  |
|  | data distribution | stato:0000161 |  |
|  | statistical sampling | stato:0000502 |  |
|  | outlier | stato:0000036 |  |
|  | primary predictor | od-attest:000015 | OD- |
|  | secondary predictor | od-attest:000016 |  |
|  | demographic distribution | od-attest:000093 | ATTEST<sup>6</sup> |
|  | outcome variable data source | od-attest:000019 |  |
|  | predictor data source | od-attest:000094 |  |
|  | cohort criteria | od-attest:000008 |  |
|  | descriptive statistic | od-attest:000012 |  |
|  | missing rate | od-attest:000068 |  |
|  | data source time coverage | od-attest:000023 |  |
|  | data source geographic coverage | od-attest:000024 |  |
|  | sources of bias | od-attest:000051 |  |
|  | data integration | od-attest:000052 |  |
|  | data extraction | od-attest:000054 |  |
|  | data cleaning | od-attest:000055 |  |
|  | integration strategy | od-attest:000056 |  |
|  | integrate variables from same level | od-attest:000057 |  |
|  | integrate variables from different levels | od-attest:000058 |  |
|  | creation of additional elements | od-attest:000059 |  |
|  | integration algorithm | od-attest:000060 |  |
|  | validation strategy | od-attest:000068 |  |
|  | integrated variable | od-attest:000096 |  |
| Properties | is determined by | od-attest:000097 | OD- |
|  | has rationale | od-attest:000098 |  |
|  | has objective | od-attest:000099 | ATTEST |
|  | has data source | od-attest:000100 |  |
|  | has cohort criteria | od-attest:000101 |  |
|  | has demographic distribution | od-attest:000102 |  |
|  | has sources of bias | od-attest:000103 |  |
|  | has controlled variable | od-attest:000104 |  |
|  | has independent variable | od-attest:000105 |  |
|  | has dependent variable | od-attest:000106 |  |
|  | has data type | od-attest:000107 |  |
|  | has descriptive statistics | od-attest:000108 |  |
|  | has NIMHD level | od-attest:000109 |  |
|  | has NIMHD domain | od-attest:000110 |  |
|  | has data collection approach | od-attest:000111 |  |
|  | has sample size | od-attest:000112 |  |
|  | has missing data | od-attest:000113 |  |
|  | has data integration | od-attest:000114 |  |
|  | has data processing | od-attest:000115 |  |
|  | has data validation | od-attest:000116 |  |
|  | has integration strategy | od-attest:000117 |  |
|  | extracted from | od-attest:000118 |  |
|  | has description | od-attest:000119 |  |
|  | has time coverage | od-attest:000120 |  |
|  | has geographic coverage | od-attest:000121 |  |
| <sup>a</sup>Prefix:<br>iao: <http://purl.obolibrary.org/obo/IAO_><br>obi: <http://purl.obolibrary.org/obo/OBI_><br>ncit: <http://ncicb.nci.nih.gov/xml/owl/EVS/Thesaurus.owl#><br>sio: <http://purl.obolibrary.org/obo/SIO_><br>stato: <http://purl.obolibrary.org/obo/STATO_><br>od-attest: <http://purl.obolibrary.org/obo/OD-ATTEST/><br><sup>1</sup> Information Artifact Ontology<br><sup>2</sup> Ontology for Biomedical Investigations<br><sup>3</sup> National Cancer Institute Thesaurus<br><sup>4</sup> Statistics Ontology<br><sup>5</sup> Semanticscience Integrated Ontology<br><sup>6</sup> Ontology for the Documentation of Variable and Data Source Selection and Integration Process |  |  |  |

### An OD-ATTEST-annotated report generated based on a mIDA case study following the reporting guideline

We annotated two of our previously published mIDA case studies: (1) one study that explored the impact of the relationships among socioeconomic status, individual smoking status, and community-level smoking rate on pharyngeal cancer survival [16], and (2) another study that created a semantic data integration framework to pool multi-level RFs from heterogenous data sources to support mIDA [25]. ***Table 4*** is the filled ATTEST checklist for the two studies. ***Figure 4*** shows a snippet of the ontology annotated variable and data source selection and integration process for the second study [25], while the corresponding semantic triples in RDF format using Turtle syntax is shown in ***Table 5***. The items: RF variables, data sources, and data integration steps and their relationships are explicitly standardized and modeled using the classes and properties from OD-ATTEST.

**Figure 4.**
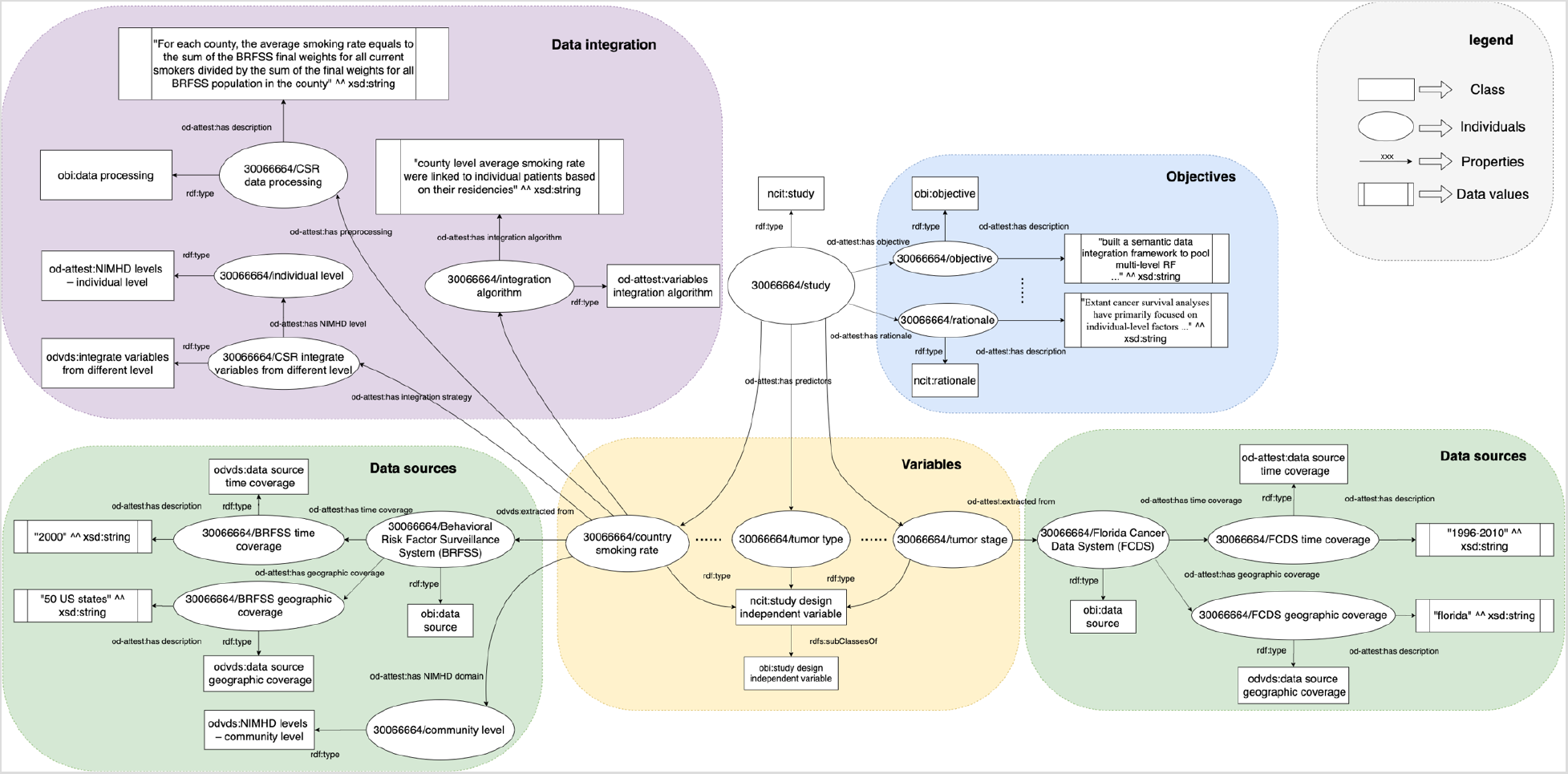
An OD-ATTEST-annotated report generated based on a mIDA case study.

**Table 4.**
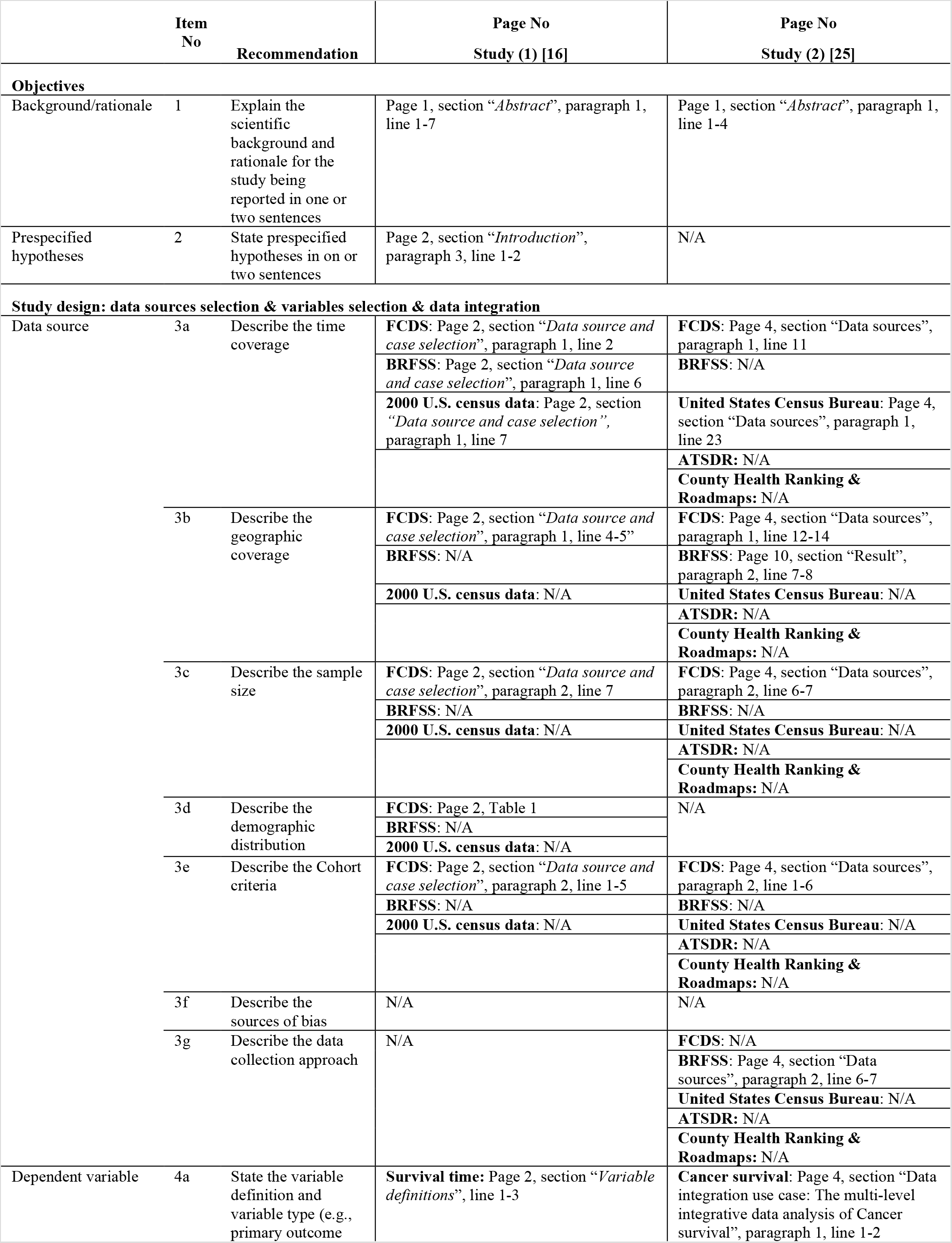
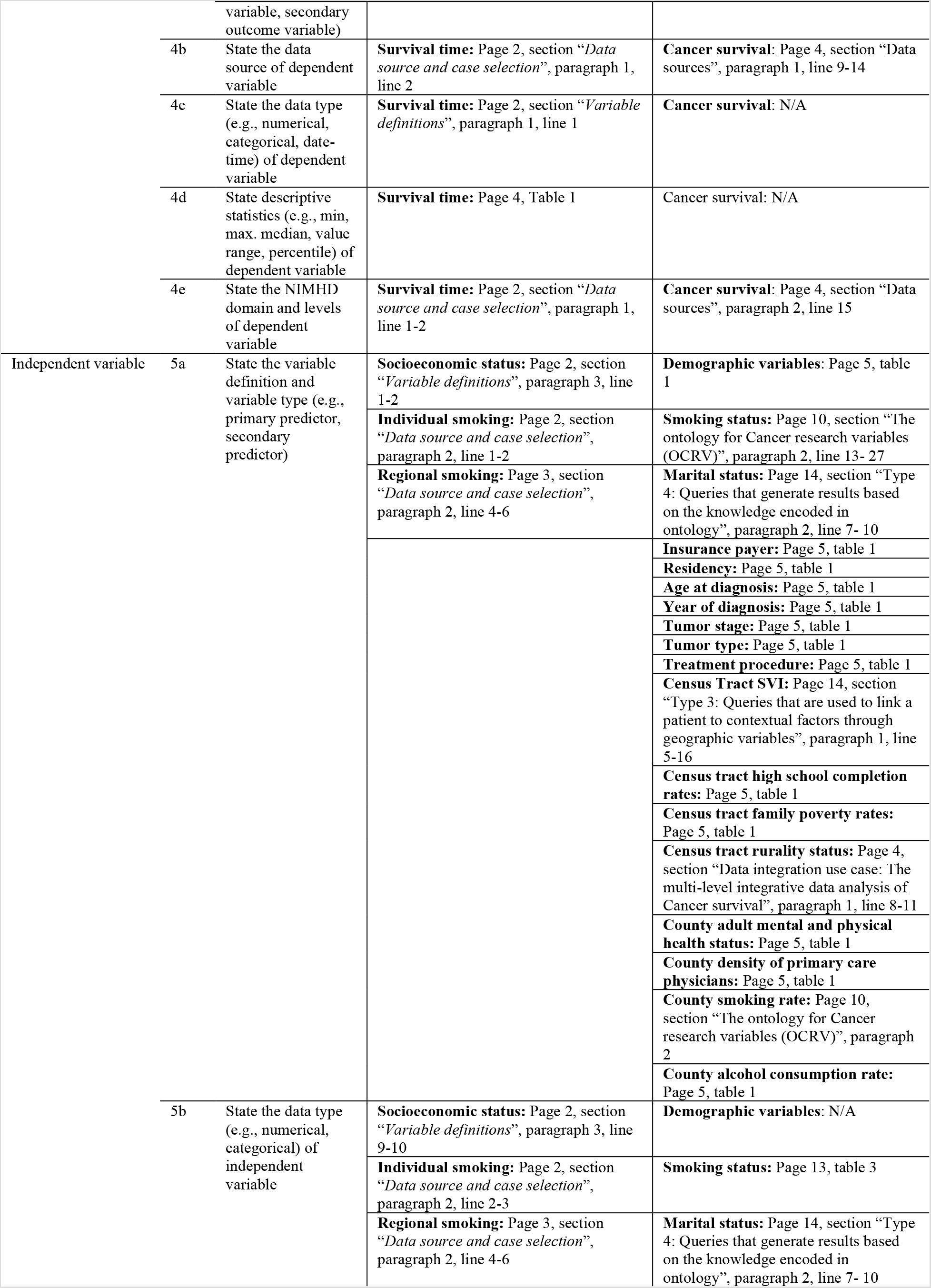
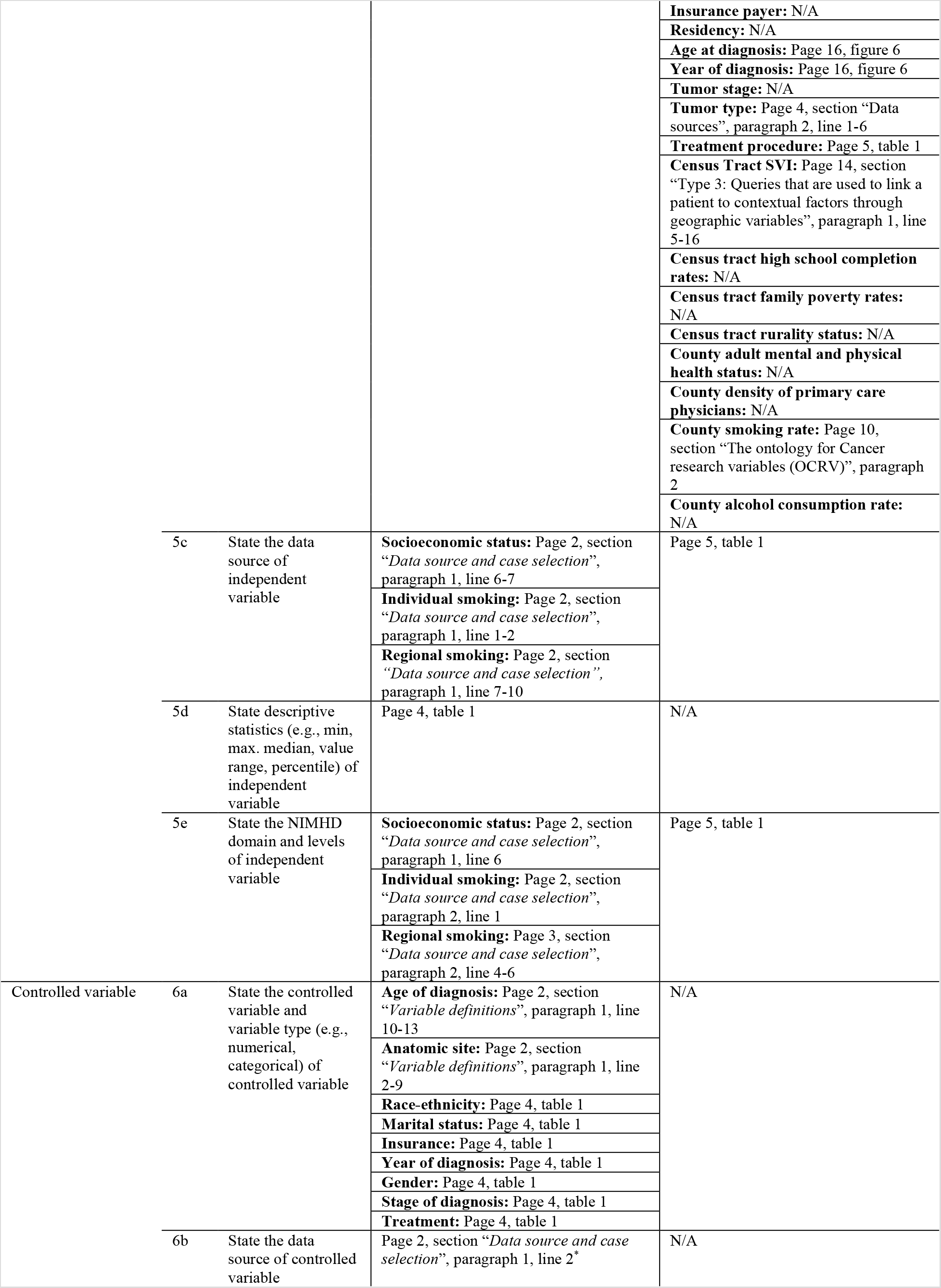
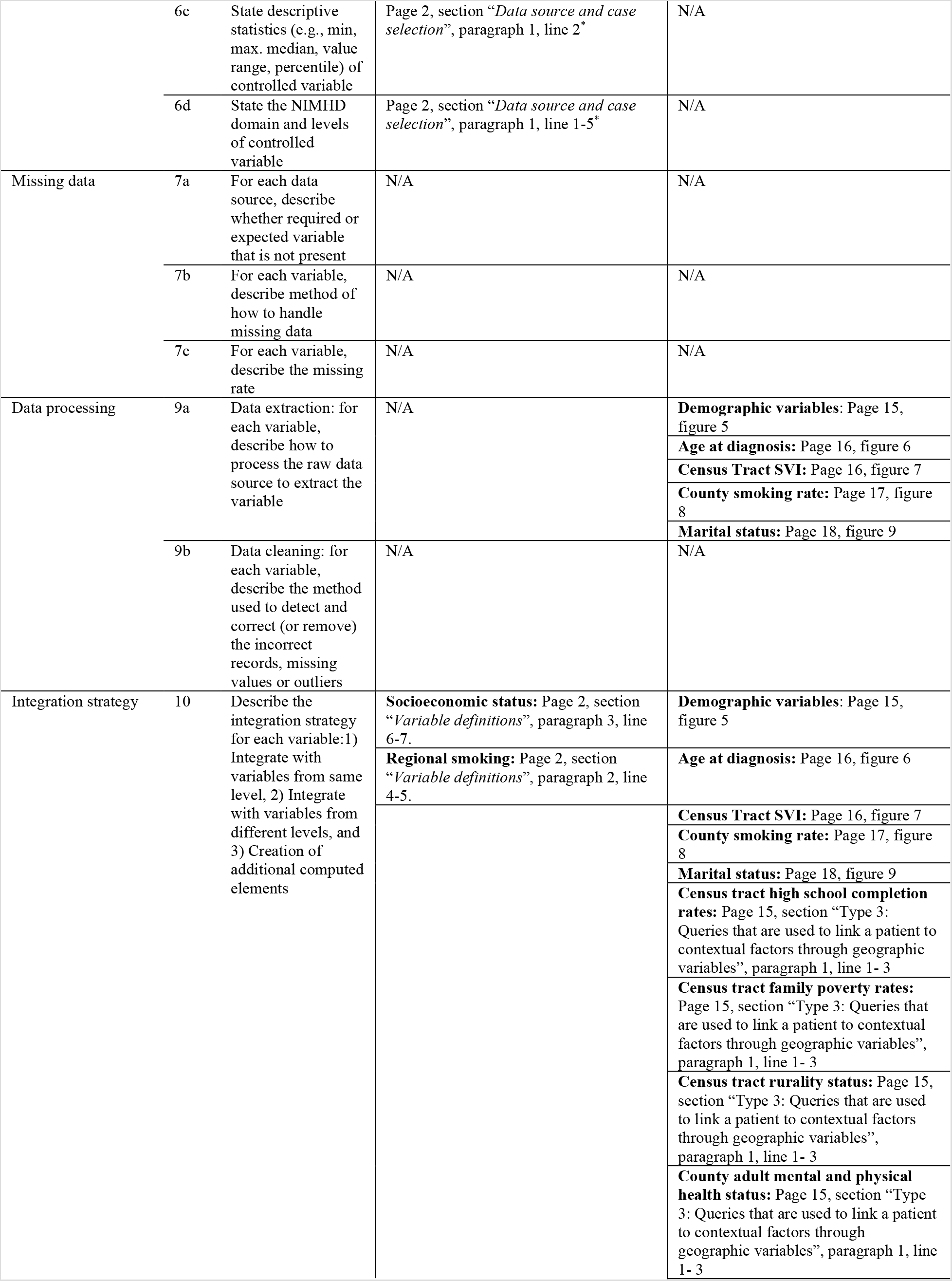
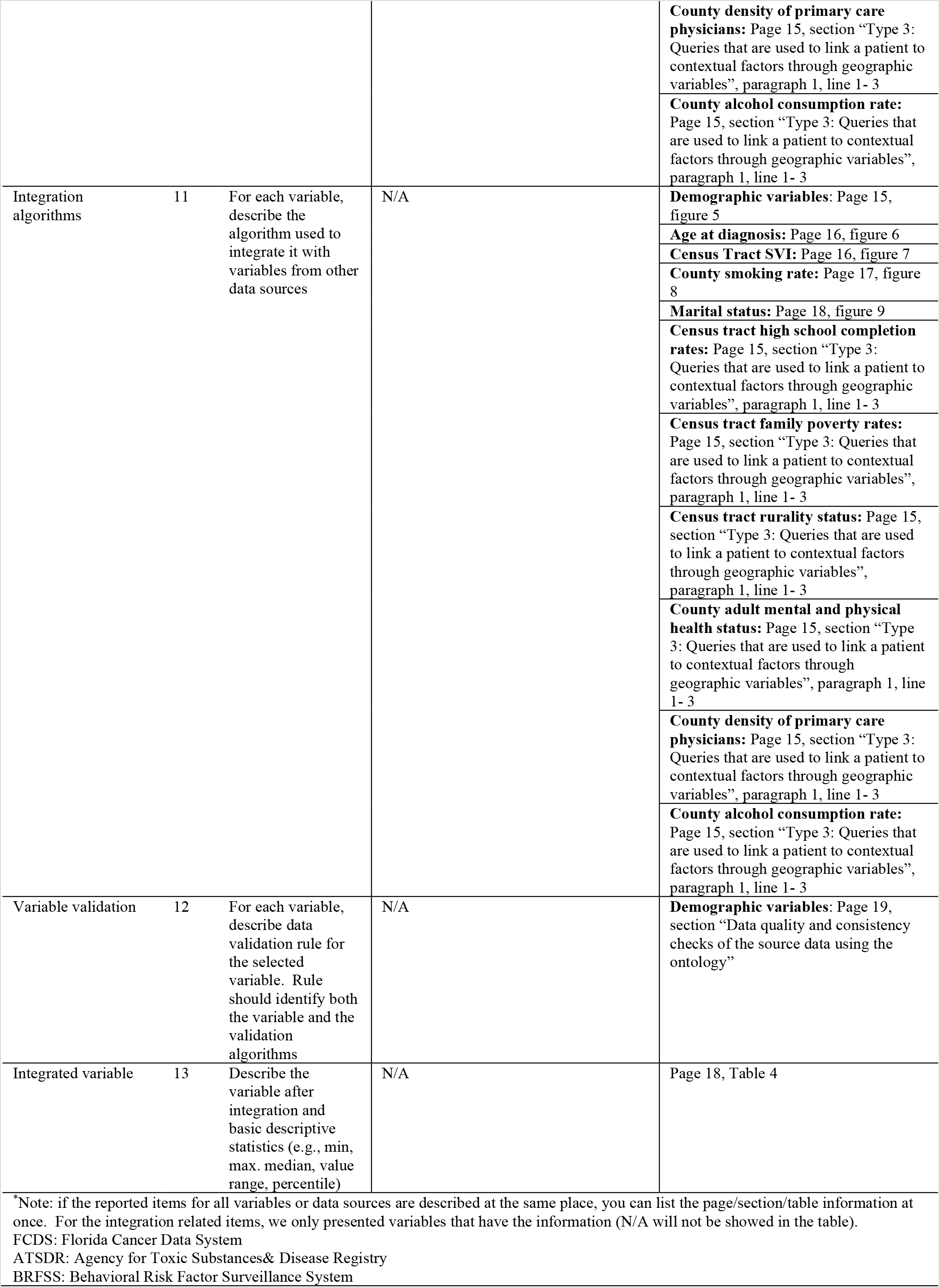
An example of two previous mIDA case studies annotated using ATTEST checklist.

**Table 5.**
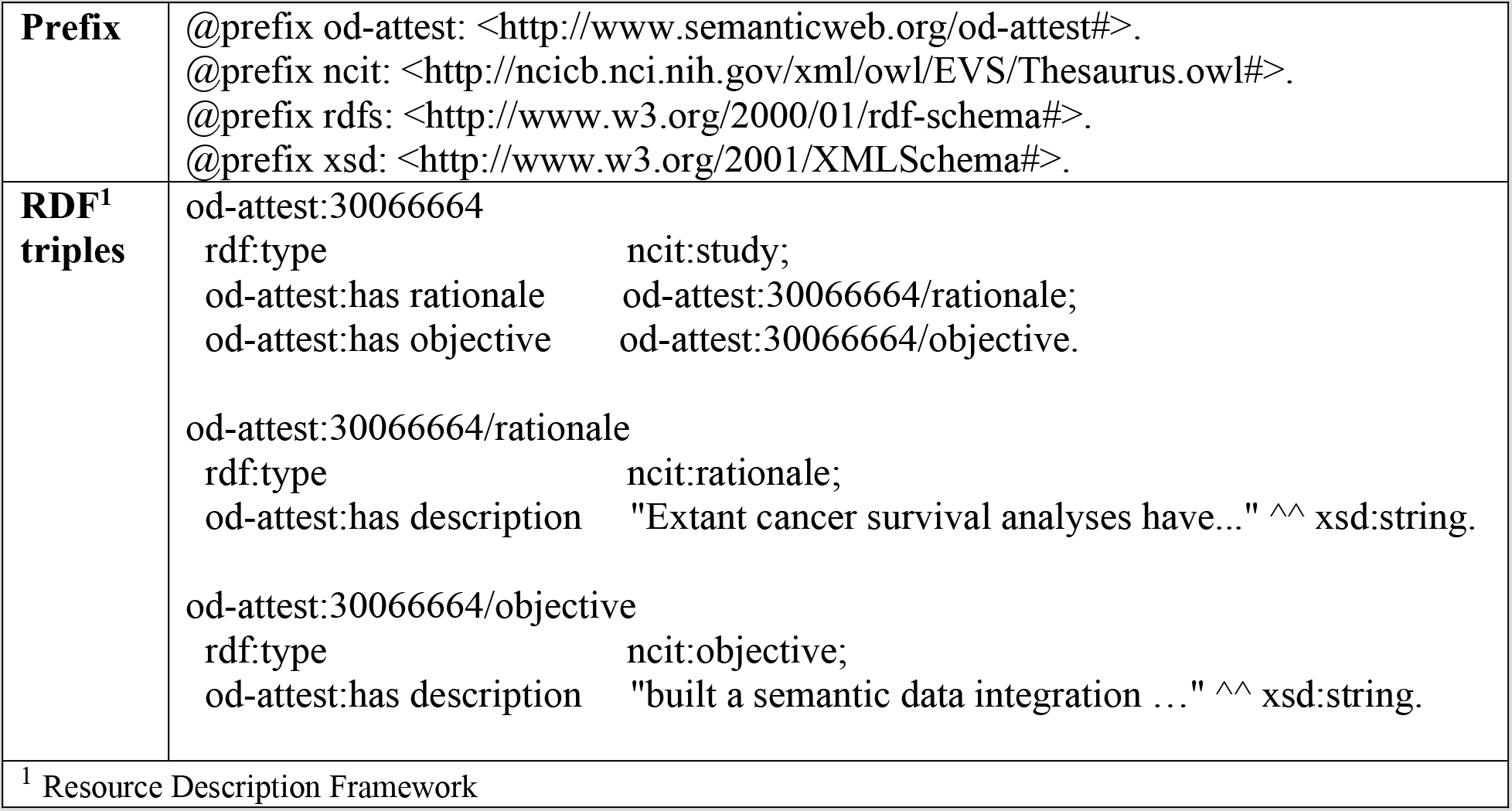
An example of annotated semantic triples represented in RDF format using Turtle syntax.

|  |  |
| --- | --- |
| **Prefix** | @prefix od-attest: < http://www.semanticweb.org/od-attest# >.<br>@prefix ncit: < http://ncicb.nci.nih.gov/xml/owl/EVS/Thesaurus.owl# >.<br>@prefix rdfs: < http://www.w3.org/2000/01/rdf-schema# >.<br>@prefix xsd: < http://www.w3.org/2001/XMLSchema# >. |
| **RDF<sup>1</sup> triples** | od-attest:30066664<br>rdf:type ncit:study;<br>od-attest:has rationale od-attest:30066664/rationale;<br>od-attest:has objective od-attest:30066664/objective.<br>od-attest:30066664/rationale<br>rdf:type ncit:rationale;<br>od-attest:has description "Extant cancer survival analyses have..." ^^ xsd:string.<br>od-attest:30066664/objective<br>rdf:type ncit:objective;<br>od-attest:has description "built a semantic data integration ..." ^^ xsd:string. |

<sup>1</sup> Resource Description Framework

## DISCUSSION

In this study, we first developed a reporting guideline, ATTEST, to provide a theory-driven approach to guide the RF variable and data source selection and integration process in cancer outcomes research. We then proposed an ontology-based approach to annotate the items in our reporting guideline so that information relevant to variables, data sources and data integration in mIDA studies can be explicitly documented. To develop the reporting guideline, we conducted a systematic search to identify useful reporting items to improve our selection and data integration process. We categorized these reporting guidelines based on their reported data source domains and levels according to NIMHD framework, so that we can identify items need to be reported when selecting variables or data sources from different domains and levels. For example, when report population-level estimates (variables) [42], the information regarding the sources of bias (e.g., selection bias) need to be documented. Therefore, we updated our previous reporting guideline and added “*sources of bias*” as a reporting item when documenting data sources. This is important, because subsequent data processing steps might be needed to correct the bias. Further,

The use of NIMHD framework can also help researchers to systematically think and structure the variable and data source selection process when considering multi-level RF variables from heterogenous data sources. For example, if an investigator is considering smoking related risk factors in cancer outcomes research, following the NIMHD framework, one can start with variables in the behavioral domain and then list potential smoking related variables for each level of influences step by step, such as individual smoking status at the individual level, second hand smoke exposure at the interpersonal level, county level smoking rate at the community level, and smoking policies or laws (e.g., federal minimum age to purchase tobacco products) at the societal level. The same process can be applied to select other smoking related variables from other domains of influences. In this way, investigators can systematically think and evaluate the confounding effects and cross-level interactions among those selected variables which are usually ignored in previous cancer outcome studies using a single data source.

We provided a ATTEST checklist (1) to help researchers clearly document each step of their RF and data source selection and integration process, and (2) to improve the completeness and transparency of their mIDA studies. As shown in ***Table 4***, we used the ATTEST checklist to report two previous mIDA studies. Based on the checklist, we can easily (1) check whether these mIDA studies document required items that can help other researchers replicate their studies, and (2) compare their variables, data sources and data integration processes. As shown in ***Table 4***,, we found that there are 3 items never discussed in either of the two studies including “*sources of bias”*, “*missing data”* for selected variables, and “*data cleaning*” (i.e., method used to detect and correct or remove the incorrect records, missing values or outliers). All three items are relevant to data quality issues, where rarely being discussed or documented in these mIDA studies or even more broadly in cancer outcomes research. Nevertheless, data quality issues such as missing data can dramatically affect the results of the cancer outcomes research (e.g., in cancer survival prediction) [60]. Comparing the two case mIDA studies, the data integration process was not well-documented in the first study [16], where most of the items relevant to data integration are blank; while, in the other study [25], the processes about data processing, data integration, and data validation were all clearly documented according to the ATTEST checklist. Therefore, using this checklist, one can improve the completeness of their documentation on the selection and integration process as shown in ***Table 4***.

The OD-ATTEST ontology provides a way to standardize the documentation of the mIDA study process from variable and data source selection to data integration. Also, the ontology-based annotations of the report is beneficial because it provides an initial step towards a report that is not only readable and understandable by human but also potentially executable by machines. After transforming these annotations into semantic triples, the report can be stored into a knowledge base and represented as knowledge graphs (***Figure 4***) to facilitate examination and analysis of these mIDA reports, enabling robust sharing and comparison of different mIDA studies.

### Limitations and future work

Most of the reporting guidelines we reviewed from the EQUATOR network have limited information on how to document the data integration process, indicating a significant gap in existing practice. Nevertheless, we were able to summarize the key elements need to be reported for the integration process based on 3 existing guidelines and our own previous experience on semantic data integration case studies. As a future study, one shall conduct a systematic review on data integration literatures to summarize relevant reporting items to improve the reporting guideline. Meanwhile, we will conduct a yearly review of existing reporting guidelines following the reviewing process discussed in ***Figure 1*** to identify new reporting items of interest and keep our framework up to date. Further, beyond standardized reporting, our ultimate goal is to let computers understand the ontology-annotated report (in RDF triples) regarding (1) how different variables are defined and represented and (2) how different variables are selected and integrated, so that machines can automatically repeat these processes and generate integrated dataset based on an executable ontology-annotated report. For variable definition and representation, it is important to recognize and being interoperable with existing data standards and common data models (CDM) such as those that standardized exchanging of EHRs data including the national Patient-Centered Clinical Research Network (PCORnet) CDM, the Observational Medical Outcomes Partnership (OMOP) from the Observational Health Data Sciences and Informatics (OHDSI) network, and the uprising Fast Healthcare Interoperability Resources (FHIR) protocol adopted by major EHR system vendors. Developing the ontology against these CDMs that have already standardized existing data resources would be critical to assure the generalizability of our framework. Nevertheless, for modeling the variable selection and integration processes as shown in ***Figure 4***, more fine-grained information regarding the variables, data sources and the integration process are currently documented as free-text descriptions. We face challenges in transforming these “free-text” information into executable algorithms (e.g., a data processing step that calculates BMI using weight and height). Such information is related to the concept of data provenance—“*a type of metadata, concerned with the history of data, its origin and changes made to it*” [61]. The importance of data provenance is widely recognized, especially for study reproducibility and replicability. More than one-half of the systematic efforts to reproduce computational results across different fields have failed, mainly due to insufficient detail on digital artifacts, such as data, code, and computational workflow [62]. However, descriptions of data provenance are often neglected or inadequate in scientific literature due to the lack of a tractable, easily operated approach with supporting tools. Future studies that focus on the development of easy-to-use tools with a standardized framework to persist end-to-end data provenance with high integrity including intermediate processes and data products are urgently needed. Further, future developments of tools and platforms to automate the documentation process, where the data elements and associated information (e.g., levels and domains) are also automatically annotated with the standardized ontology are warranted.

## CONCLUSIONS

In this paper, we have proposed and developed an ontology-based reporting guideline solving some key challenges in current mIDA studies for cancer outcomes research, through providing (1) a theory-driven guidance for multi-level and multi-domain RF variable and data source selection; and (2) a standardized documentation of the data selection and integration processes powered by an ontology, thus a way to enable sharing of mIDA study reports among researchers.

## Data Availability

All the data needed are included in the manuscript.

## List of abbreviations

ACS: American Cancer Society
BRFSS: Behavioral Risk Factor Surveillance System
EQUATOR: Enhancing the QUAlity and Transparency Of health Research
FCDS: Florida Cancer Data System mIDA Multi-level Integrative Data Analysis
NIH: National Institute of Health
NIMHD: Minority Health and Health Disparities
OD-ATTEST: Ontology for the Documentation of Variable and Data Source Selection and Integration Process
RF: Risk Factor
US: United States
RUCA: Rural-Urban Commuting Area
NCHS: National Center for Health Statistics
BFO: Basic Formal Ontology
NCBO: National Center for Biomedical Ontology
RDF: Resource Description Framework
GRIPS: Genetic RIsk Prediction Studies
COHERE: Checklist for One Health Epidemiological Reporting of Evidence EHR Electronic Health Records
OBI: Ontology for Biomedical Investigations
IAO: Information Artifact Ontology
NCIt: National Cancer Institute Thesaurus
STATO: Statistics Ontology
SIO: Semanticscience Integrated Ontology
CDM: Common Data Model
PCORnet: The national Patient-Centered Clinical Research Network

## Declarations

### Ethics approval and consent to participate

Not applicable

### Consent to publish

Not applicable

### Availability of data and materials

The reviewed reporting guidelines are available in the public: Enhancing the QUAlity and Transparency Of health Research (EQUATOR) network (https://www.equator-network.org/reporting-guidelines/).

### Competing interests

The authors declare that they have no competing interests.

### Funding

This study was supported in part by the National Institute of Health (NIH) awards UL1TR001427 and R01CA246418 and Patient-Centered Outcomes Research Institute (PCORI) award ME-2018C3-14754. The content is solely the responsibility of the authors and does not necessarily represent the official views of the NIH or PCORI.

### Authors’ contributions

The work presented here was carried out in collaboration among all authors. YG and JB designed the study. YG, QL and HZ were involved in acquisition of the data and review of existing reporting guidelines. HZ wrote the initial draft of the manuscript with substantial support from YG and JB. YG and JB provided expert opinion during the ontology curation process and guided the design of the ontology. All authors provided critical feedback on the study design, reviewed and edited the manuscript. All authors have read and approved the final manuscript.

## Acknowledgements

None

## Notes

### Competing Interest Statement

The authors have declared no competing interest.

### Author Declarations

Our study doesn't require IRB approval.

